# Huntingtin gene CAG repeat size in patients with Lynch syndrome

**DOI:** 10.1101/2022.05.28.22275723

**Authors:** Karin Dalene Skarping, Åsa Petersén, Samuel Gebre-Medhin

## Abstract

Patients with Lynch syndrome (LS) are prone to cancer due to heterozygous germline pathogenic variants in genes encoding DNA mismatch repair proteins MLH1, MSH2, MSH6 and PMS2. LS cancer cells exhibit deficient DNA mismatch repair and microsatellite instability due somatic inactivation of the second copy of the affected gene. To study microsatellite characteristics in non-neoplastic cells in LS we determined CAG repeat size in the huntingtin gene (*HTT*) microsatellite in lymphocyte DNA from LS patients with germline pathogenic variants in *MLH1* (n = 11), *MSH2* (n = 9), *MSH6* (n = 7) and non-LS controls (n=19). Mean repeat size in LS was 19,55 CAG (*MLH1*), 19,39 CAG (*MSH2*), 18.07 CAG (*MSH6*), respectively compared to 18,42 CAG in controls. Standard deviation for CAG repeat size in LS was 4,183 CAG (*MLH1*), 5,089 CAG (*MSH2*), 3,075 CAG (*MSH6*), respectively, compared to 3,342 CAG in controls. Peak CAG repeat size in LS was 32 CAG (*MLH1*), 32 CAG (*MSH2*), 24 CAG (*MSH6*), respectively compared to 27 CAG in controls. Collectively, our data indicate that *HTT* CAG repeat size tends to be larger and more variable in individuals with LS caused by pathogenic variants in *MLH1* and *MSH2*.

## Introduction

Lynch syndrome (LS) is a multiorgan cancer predisposition syndrome caused by germline heterozygous pathogenic variants (PV) in the mismatch repair (MMR) genes *MLH1, MSH2, MSH6* or *PMS2* ^1^. LS cancers exhibit deficient MMR (dMMR) and microsatellite instability (MSI) due to somatic inactivation of the remaining allele of the affected MMR gene ^1^. In the vast majority of patients, the underlying germline PV is inherited, and as a consequence there is often an accumulation of LS cancers (predominantly colon cancer and uterus cancer) in the affected family branch. Average age at onset is lower for LS cancers than for sporadic cancers. In addition, anticipation, i.e. lower age at disease onset in subsequent generations has been reported in LS ^2,3^ although the phenomenon has been questioned in the absence of mechanistic explanation or explained by a birth-cohort bias ^4,5^. Yet, as the MMR system is ubiquitous to maintain nuclear genome stability ^6^ dysfunctional MMR alleles could conceivably contribute to an increased mutational load including in germ cells and hence acquired genetic aberrations could be transmitted to the next generation. Indeed, the state of haploidy in gametes implies that half of the germ cells in patients with LS have no functional copy of the affected MMR gene and could therefore be subject to dMMR. However, a recent whole genome sequencing (WGS) effort revealed no evidence for altered mutational load in non-neoplastic tissue in LS patients ^7^. Since WGS technologies still lack sufficient resolution in regions with short tandem repeats (STR), i.e. in regions with microsatellites, we herein have used a PCR-based clinical grade high-resolution DNA fragment analysis to determine the range of CAG repeats in the huntingtin gene (*HTT*) microsatellite in patients with LS and matched controls.

## Materials and Methods

Genomic DNA extracted from blood samples from patients investigated for non-polyposis hereditary colorectal cancer (CRC) in the Southern health care region in Sweden during 1997-2012 and shown to have either a dMMR colorectal cancer (CRC) and a PV diagnostic for LS or a MMR proficient CRC (referred to as controls in the present study) were retrieved from the Skåne university hospital biobank and anonymized. LS patients with PV in *PMS2* were not included in this study for reasons of integrity as they were too few cases in the clinical registry. *HTT* CAG repeat size estimation was performed using PCR amplification and capillary electrophoresis fragment analysis with a validated accuracy of ± 1 CAG repeat for alleles with < 45 repetitions and ± 3 CAG repetitions for alleles with 45 or more repeats as described ^8^. *HTT* was chosen as it contains a well-characterized STR known to contain pathogenic CAG repeat expansions causative for Huntington disease (HD) and since the *HTT* CAG pathogenic repeat is thought to further expand somatically in an MMR-dependent manner ^9^.

### Statistical analyses

*HTT* CAG repeat values were converted to integers according to clinical genetic laboratory diagnostic routines for Huntington disease ^8^. The CAG repeat size estimation error +/-1 was excluded from statistical calculations. Due to limited number of patients and incomplete normal distribution of histogram data, Mann-Whitney U and Kruskal-Wallis tests were used. CAG repeat size was analyzed either as unpaired values (independent alleles) or as paired (i.e. as the sum of CAG repeats for both *HTT* alleles in each patient). A CAG repeat size of of 18,42 triplets was set at baseline in accordance with values from patient controls in the present study and in line with published data from 7379 unselected individuals in Sweden (Sundblom et al., 2020). SPSS Statistics for Windows (SPSS Inc., Chicago, Ill., USA) software was used. *P-*values < 0.05 were considered significant.

### Ethical approvals

Approvals and decisions were received from The Regional Ethical Review Board in Lund (application no. 2013/468) and from the Swedish Ethical Review Agency (application no. 2019-02312 and application no. 2021-06254-02).

## Results

CAG repeat size was determined for both *HTT* alleles in LS patients with PV in *MLH1* (n = 11), *MSH2* (n = 9), *MSH6* (n = 7) and in control patients (n=19) (Table 1, Figure 1). Mean CAG repeat size for unpaired *HTT* alleles in the LS subgroups was 19,55 CAG (*MLH1*), 19,39 CAG (*MSH2*), 18,07 CAG (*MSH6*), respectively compared to 18,42 CAG in controls (Figure 2). Standard deviation (SD) for CAG repeat size for unpaired *HTT* alleles in the LS subgroups was 4,183 CAG (*MLH1*), 5,089 CAG (*MSH2*), 3,075 CAG (*MSH6*), respectively, compared to 3,342 CAG in controls (Figure 2). SD for the sum of CAG repeat size for paired *HTT* alleles in LS subgroups was 6,640 CAG (*MLH1*), 6,667 CAG (*MSH2*), 3,761 CAG (*MSH6*), respectively, compared to 4,375 CAG in controls (Figure 3). Peak CAG repeat size in LS subgroups was 32 CAG (*MLH1*), 32 CAG (*MSH2*), 24 CAG (*MSH6*), respectively compared to 27 CAG in controls (Table 1; Figure 1). Mean sum of CAG repeat size for paired *HTT* alleles in LS subgroups was 39,09 CAG (*MLH1*), 38,78 CAG (*MSH2*), 36,14 CAG (*MSH6*), respectively compared to 36,84 CAG in controls (Figure 3). Differences observed between the LS subgroups or between LS subgroups and controls were not statistically significant (data not shown).

**Table 1.** Summary of *HTT* CAG repeat size data for all patients in the study. Patient ID includes Lynch syndrome genetic subcategory (i.e. pathogenic variant in *MLH1, MSH2* or *MSH6*, respectively) and controls (bottom). Clinical value refers to the CAG repeat size that would have been reported in a clinical laboratory routine. Column headed Sum denotes sum of CAG repeat size for allele 1 and allele. Column headed Difference denotes difference in CAG repeat size between allele 2 and allele 1.

| CAG repeat size (clinical value) |  |  |  |  |
| --- | --- | --- | --- | --- |
| Patient ID | Allele 1 | Allele 2 | Sum | Difference |
| 1 MLH1 | 16,7 (17) | 16,7 (17) | 34 | 0 |
| 2 MLH1 | 18,6867 (19) | 18,6867 (19) | 38 | 0 |
| 3 MLH1 | 16,6967 (17) | 17,7467 (18) | 35 | 1 |
| 4 MLH1 | 17,7067 (18) | 18,7033 (19) | 37 | 1 |
| 5 MLH1 | 16,64 (17) | 17,7467 (18) | 35 | 1 |
| 6 MLH1 | 16,6567 (17) | 18,68 (19) | 36 | 2 |
| 7 MLH1 | 12,51 (13) | 17,7467 (18) | 31 | 5 |
| 8 MLH1 | 17,6667 (18) | 23,6367 (24) | 42 | 6 |
| 9 MLH1 | 17,7467 (18) | 25,7267 (26) | 44 | 8 |
| 10 MLH1 | 16,7 (17) | 25,75 (26) | 43 | 9 |
| 11 MLH1 | 22,7033 (23) | 32,07 (32) | 55 | 9 |
| 1 MSH2 | 16,7 (17) | 16,7 (17) | 34 | 0 |
| 2 MSH2 | 17,7867 (18) | 19,7533 (20) | 38 | 2 |
| 3 MSH2 | 17,79 (18) | 21,7367 (22) | 40 | 4 |
| 4 MSH2 | 16,59 (17) | 21,6067 (22) | 39 | 5 |
| 5 MSH2 | 11,4467 (11) | 15,6767 (16) | 27 | 5 |
| 6 MSH2 | 17,79 (18) | 23,7733 (24) | 42 | 6 |
| 7 MSH2 | 16,6333 (17) | 25,71 (26) | 43 | 9 |
| 8 MSH2 | 11,3033 (11) | 23,64 (24) | 35 | 13 |
| 9 MSH2 | 18,7467 (19) | 32,0533 (32) | 51 | 13 |
| 1 MSH6 | 17,7467 (18) | 17,7467 (18) | 36 | 0 |
| 2 MSH6 | 16,6767 (17) | 17,7467 (18) | 35 | 1 |
| 3 MSH6 | 18,6967 (19) | 19,6533 (20) | 39 | 1 |
| 4 MSH6 | 14,5767 (15) | 16,6967 (17) | 32 | 2 |
| 5 MSH6 | 13,4067 (13) | 17,6233 (18) | 31 | 5 |
| 6 MSH6 | 16,7 (17) | 23,6833 (24) | 41 | 7 |
| 7 MSH6 | 14,5733 (15) | 23,68 (24) | 39 | 9 |
| 1 Control | 20,67 (21) | 20,67 (21) | 42 | 0 |
| 2 Control | 14,5033 (15) | 15,5567 (16) | 31 | 1 |
| 3 Control | 16,6533 (17) | 17,7067 (18) | 35 | 1 |
| 4 Control | 16,6167 (17) | 17,6667 (18) | 35 | 1 |
| 5 Control | 17,7067 (18) | 18,69 (19) | 37 | 1 |
| 6 Control | 18,69 (19) | 19,6767 (20) | 39 | 1 |
| 7 Control | 15,6033 (16) | 17,7067 (18) | 34 | 2 |
| 8 Control | 16,6567 (17) | 18,6867 (19) | 36 | 2 |
| 9 Control | 14,5 (15) | 17,6667 (18) | 33 | 3 |
| 10 Control | 16,65 (17) | 19,68 (20) | 37 | 3 |
| 11 Control | 15,6033 (16) | 18,6833 (19) | 35 | 3 |
| 12 Control | 14,49 (14) | 17,6667 (18) | 32 | 4 |
| 13 Control | 17,7067 (18) | 21,59 (22) | 40 | 4 |
| 14 Control | 17,7467 (18) | 21,6767 (22) | 40 | 4 |
| 15 Control | 18,6867 (19) | 25,6633 (26) | 45 | 7 |
| 16 Control | 14,5167 (15) | 22,59 (23) | 38 | 8 |
| 17 Control | 14,5767 (15) | 23,6933 (24) | 39 | 9 |
| 18 Control | 9,2567 (9) | 18,68 (19) | 28 | 10 |
| 19 Control | 16,65 (17) | 26,6833 (27) | 44 | 10 |

**Figure 1.**
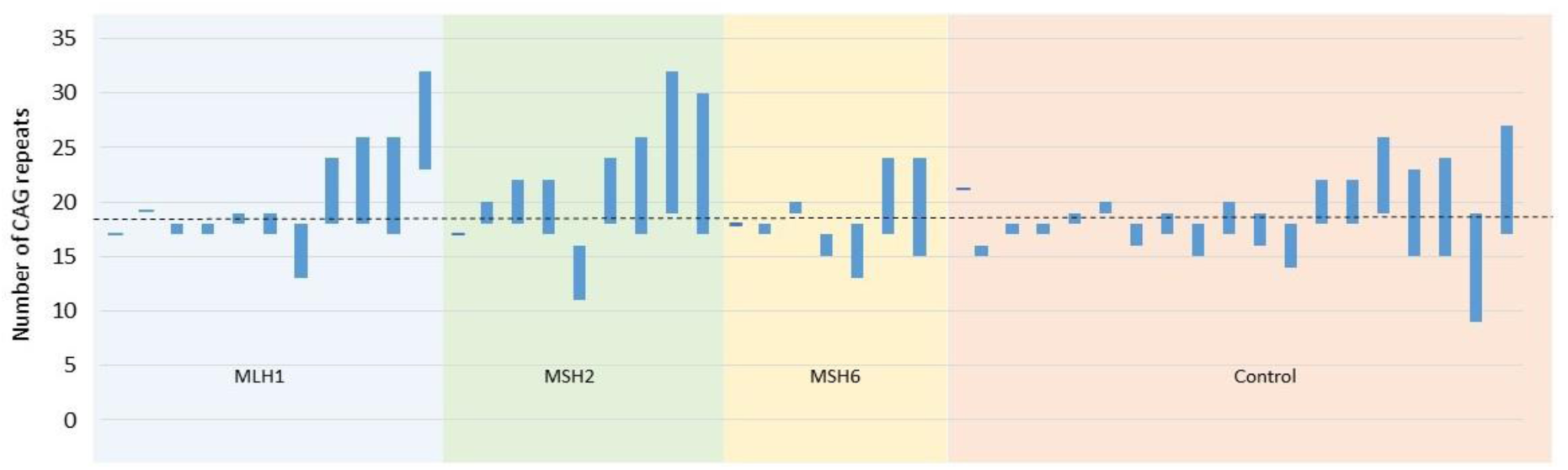
Graphical representation of *HTT* CAG repeat size in all patients. Bar representation of allele 1 (upper end of bar) and allele 2 (lower end of bar) CAG repeats size for each patient in the study. Bars are grouped for patients with Lynch syndrome due to pathogenic variants in *MLH1* (blue field), *MSH2* (green field), *MSH6* (yellow field) and control patients (red field). Mean CAG repeat size for control patients is shown (dashed line).

**Figure 2.**
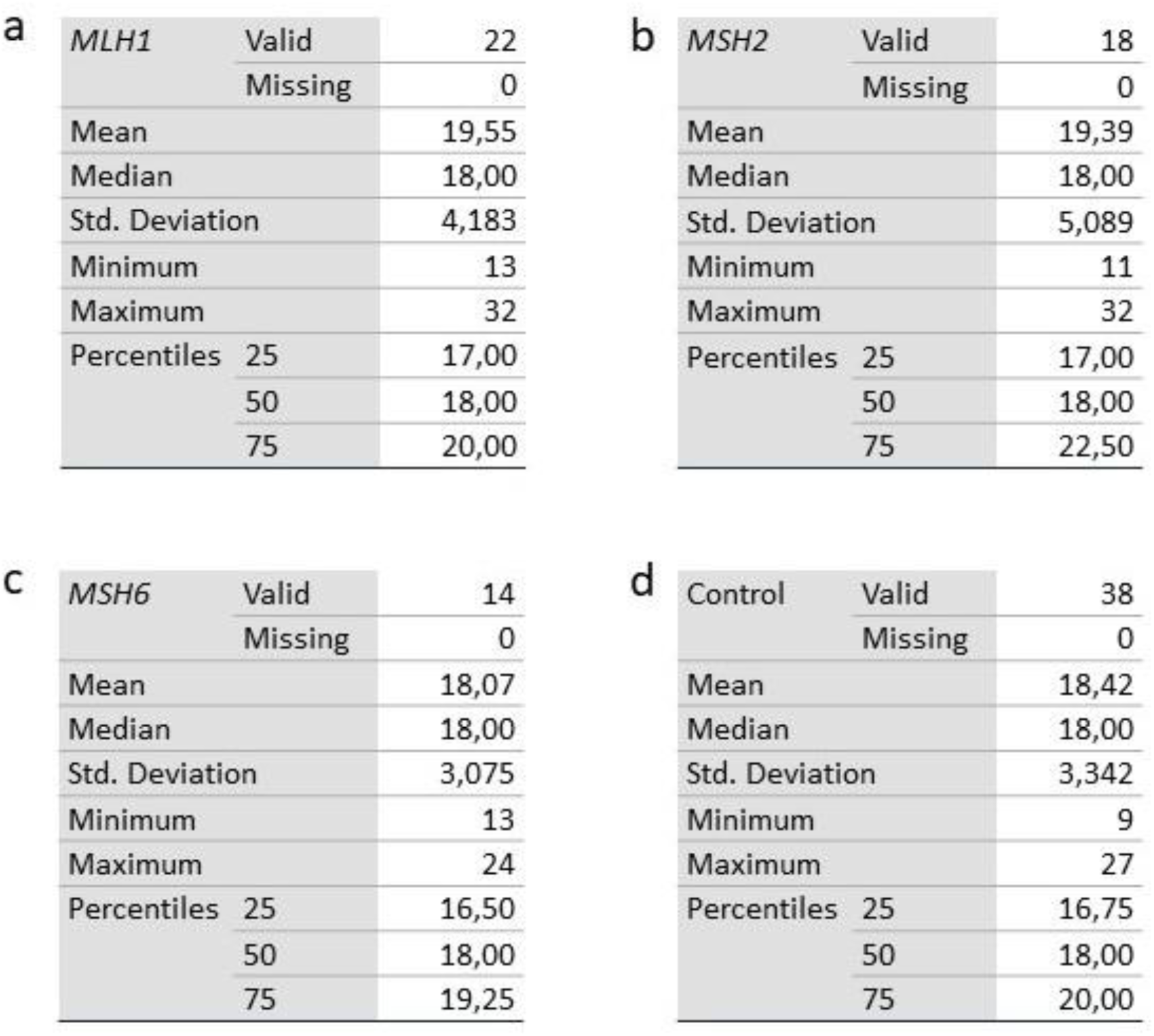
Statistics of CAG repeat size for unpaired *HTT* alleles. Statistic evaluation of CAG repeat size data for unpaired *HTT* alleles in patients with Lynch syndrome with a pathogenic variant in *MLH1* (a), *MSH2* (b), *MSH6* (c), and controls (d)

**Figure 3.**
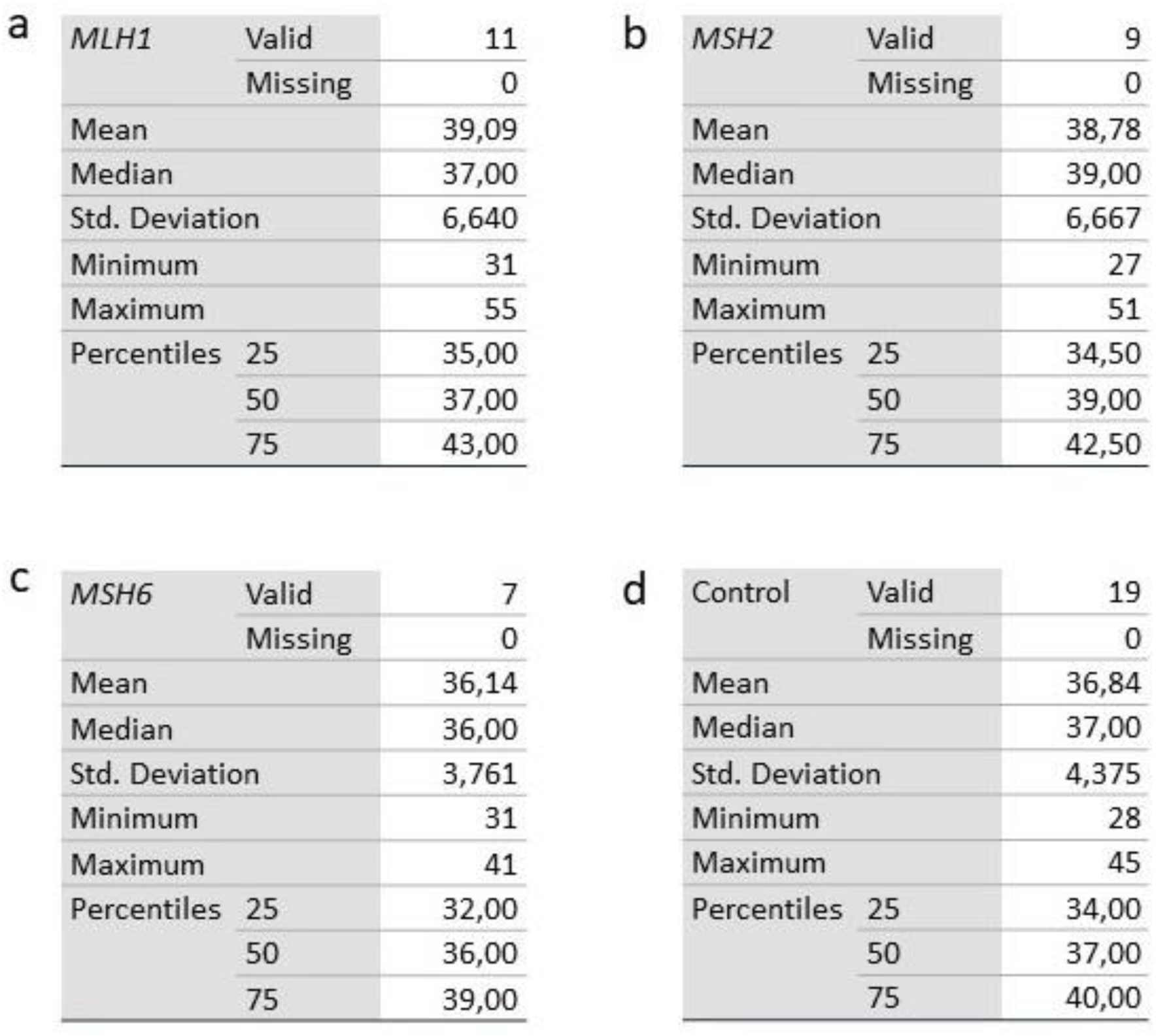
Statistics of CAG repeat size for paired *HTT* alleles. Statistic evaluation of sum of CAG repeat size, i. e. paired *HTT* alleles in patients with Lynch syndrome with a pathogenic variant in *MLH1* (a), *MSH2* (b), *MSH6* (c), and controls (d).

## Conclusion

In this study we hypothesized that heterozygosity for PV in MMR genes could have an impact on microsatellite size in non-neoplastic cells. For this purpose we studied CAG repeat size in the *HTT* microsatellite in genetic subgroups of patients with LS. We found that CAG repeat size in LS patients with PV in *MLH1* and *MSH2* tended to be larger and more variable compared to patients with a PV in *MSH6* and non-LS controls. The differences observed between the groups were however not statistical significant. The study of a larger group of patients with LS or inter-generational studies of LS family members could possibly clarify whether or not PV in MMR genes affect microsatellite length in germline DNA.

## Data Availability

All data produced in the present study are available upon reasonable request to the authors.

## Acknowledgements

We wish to thank the Department of Clinical Genetics and Pathology, Office for Medical Service for support regarding personnel, equipment and materials in this study.

